# Validation of NUTRENTO web app for teachers to assess healthy food environment practices in Mexican schools

**DOI:** 10.1101/2025.08.18.25333945

**Authors:** Jhazmín Hernández-Cabrera, Marcos Galván, Lilia Castro-Porras, Luis A. Moreno-Aznar, Guadalupe López-Rodríguez, Carolina Pérez-Ferrer, Norma Angélica Ortega-Andrade

**Affiliations:** PhD Student in Healthy Behavior Sciences, School of Health Sciences, Universidad Autónoma del Estado de Hidalgo. 42160, Pachuca, Hidalgo, México; Policy Population and Health Research Center, School of Medicine, National Autonomous University of Mexico. Ciudad Universitaria, CDMX; Center for Biomedical Research and Physiopathology in Obesity and Nutrition (CIBERObn), University of Zaragoza. Zaragoza Spain; Nutritional Epidemiology Research Group (CAEPINUT), School of Health Sciences, Universidad Autónoma del Estado de Hidalgo, Pachuca, Hidalgo, México; Population Health Research Center, Instituto Nacional de Salud Pública. Cuernavaca, México; Cuerpo Académico de Salud Emocional, School of Health Sciences, Universidad Autónoma del Estado de Hidalgo, Pachuca, Hidalgo, México; Interdisciplinary Network of Experts in School Environments in Latin America. Pachuca de Soto, Hidalgo, México

**Keywords:** food environment, validation, mobile Applications, obesity prevention, schools

## Abstract

In countries such as Mexico, where childhood obesity remains a pressing public health concern, schools are crucial environments for promoting healthy eating habits among children. Although national policies exist to improve school food environments, their implementation at the local level often lacks monitoring and support. Teachers are essential actors in translating these policies into everyday practices. However, validated tools for identifying and strengthening healthy food environments are lacking. The objective of this study was to develop and validate a progressive web application aimed at primary school teachers to identify the level of actions and elements that foster healthy school food environments in Mexico. A questionnaire based on ecosocial theory was developed and validated by expert judgment, covering seven dimensions related to healthy school food environments. A progressive web application (PWA) was developed, which included its own content management system (CMS). A total of 1462 teachers from 123 randomly selected schools completed the questionnaire using computers or mobile devices. Exploratory and confirmatory factor analysis (CFA) methods were employed to validate the scale. Results. From the CMS, 970 complete responses were retrieved for 52 content-validated items. In the EFA, there were six factors for 36 items, indicating excellent sampling adequacy (KMO> 0.8). CFA indicated excellent model fit (CFI = 0.95, TLI = 0.945, RMSEA = 0.045, SRMR = 0.059). Cronbach’s alpha values ranged from 0.80 to 0.95, confirming internal consistency. The app represents the first validated digital tool in Mexico with open access for any device. The PWA is a large-scale instrument that provides real-time data to identify teachers’ roles as mediators of the school food environment, helping prioritize and monitor interventions within schools.

## Introduction

Scientific evidence has shown that to address the growing pandemic of childhood overweight (OW) and obesity (OB) effectively in low- and middle-income countries, such as Mexico, where 36.5% of school-aged children suffer from these conditions [1], prevention and treatment strategies should be implemented to address multiple factors, considering the school food environments where children spend much of their time [2–4].

A school food environment (SFE) is defined as all spaces, infrastructure, and conditions in and around school facilities where food is available, obtained, purchased, and/or consumed (e.g., grocery stores, kiosks, canteens, food vendors, vending machines, etc.). It includes the nutritional content of these foods, as well as all available information, promotion (marketing, advertising, branding, food labels, packages, promotions, etc.), food products and their cost. A healthy SFE (HSFE) allows and encourages the school community (children, school staff, etc.) to choose foods that contribute to improved diets and greater well-being [5].

A HSFE facilitates and promotes meaningful learning in children’s healthy lifestyles, fostering and mediating interactions between the approach of the contents of nutritional food education and the practice of this knowledge within the social environment of the school [5]. On the other hand, an unhealthy SFE can contribute to the gain of adiposity in childhood, having few or no actions or elements related to the access, availability or promotion of healthy foods [6,7]. In school contexts, promoting a HSFE is a challenge, as schools are involved in numerous curricular and social activities [8]. Promoting and mediating a HSFE means addressing different areas beyond compliance with regulations, available information, food outlets, hygiene, or food guidelines. [9]

Teachers (TEs) represent one of the main mediators of the SFE, as in their role, they implement actions linked to regulations, the curriculum, and programs for the SFE. They could therefore strengthen and carry out actions that positively impact the SFE, thereby contributing to limiting the increasing prevalence of childhood OW and OB. [10–15].

In the Mexican context, the school environment and its recent system of management and regulation in terms of actions to promote healthy eating and lifestyles make it possible that, from the organization of the school community, particularly from TEs, as mediators of SFE and school leaders, can collaborate with the school social participation councils, address areas of opportunity and carry out contextualized actions in schools. Actions can be planned and implemented across various areas, ranging from linkages with public agencies related to school health, budgeting and financial oversight, infrastructure, and healthy eating, among others. In addition, in the current basic education curriculum in Mexico, there are specific themes and contents associated with the promotion of healthy eating habits[16]. This could represent a valuable opportunity to promote HSFE and contribute more effectively to child health care[17]. However, one of the challenges in Mexico is for teachers to adapt to new regulations and public policies, such as “Vive Saludable”, and to the organization and prioritization of school actions [18,19].

Owing to the increasing interest in and significance of food environments [12–14,20,21], methods have been developed for their study and analysis in different settings, evaluating their components and associations with OW and OB in childhood and school [22–26]. M-Health, in frameworks such as progressive web applications (PWAs), is an accessible and scalable option to support the implementation of school health policies. [26–29]. PWAs can be used on any internet-enabled device, facilitating real-time data collection and monitoring of school actions, even in resource-limited settings. [30]

However, regardless of the technology used, to identify an HSFE, it is necessary to have validated evaluation instruments that integrate the teacher as the main mediator of the actions to promote an HSFE [31–33]. In Mexico, there is no standardized and validated instrument that allows the immediate and large-scale identification of the dimensions and actions carried out in HFSEs. Identifying the level of actions that a school performs could be beneficial for the prioritization of actions within heterogeneous contexts [17]. There are few HSFE instruments with construct validity [31,32], so it is necessary to develop self-report instruments aimed at teachers, which are based on M-Health, to evaluate different components of the school food environment and identify areas to initiate actions, increase their level, or maintain them [33,34]. The objective of this work was to develop and validate a PWA aimed at primary school teachers to identify the level of actions and elements that promote healthy school food environments in Mexico.

## Materials and methods

### Study design

This was a cross-sectional validation study of an online instrument developed in four stages: conceptualization, content validation, web application development and construct validation (Fig. 1).

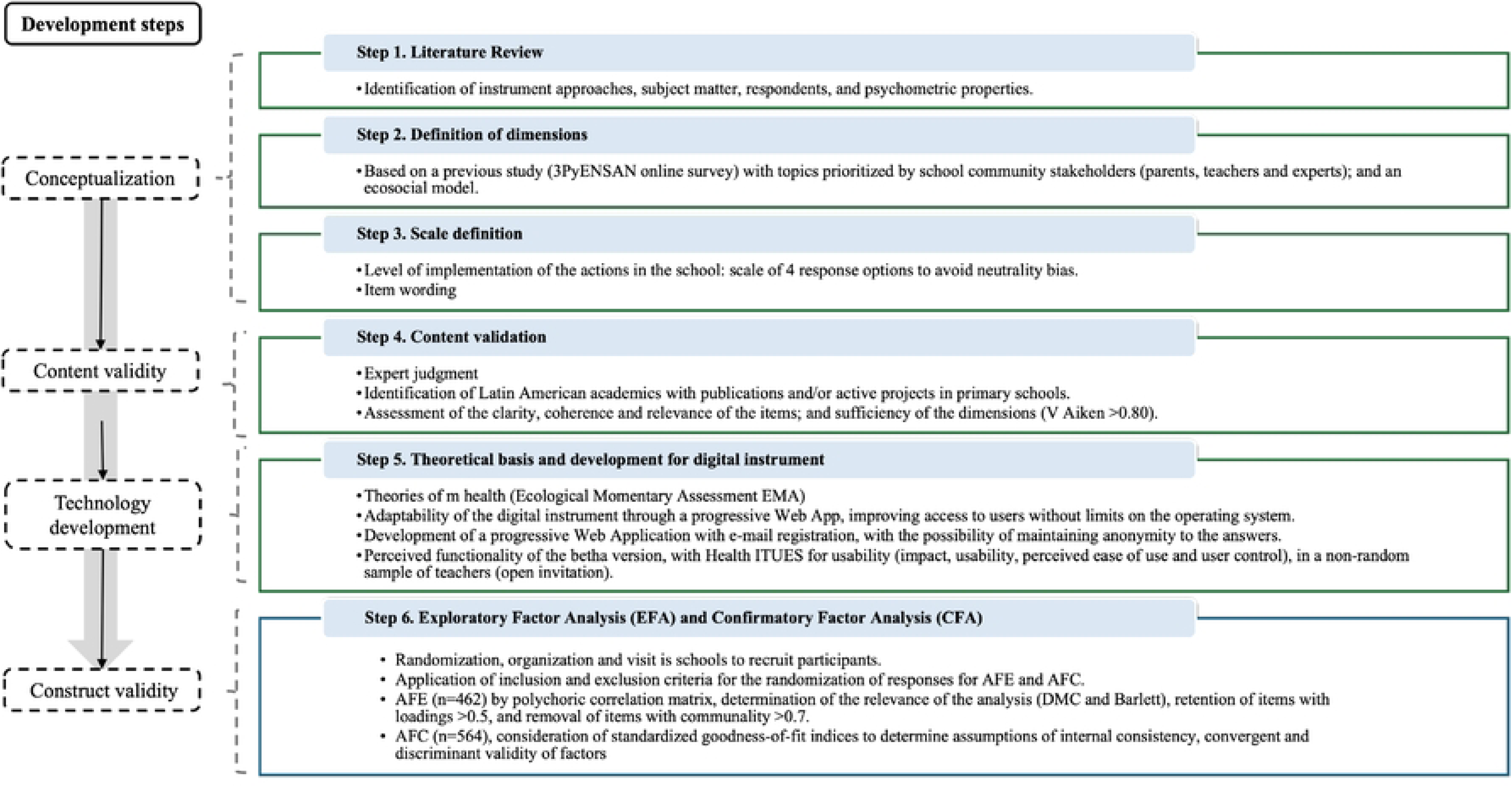
Flow chart of the stages and steps for the development and validation of the NUTRENTO Progressive Web Application.

### Conceptualization of the questionnaire

First, the literature was reviewed, identifying previous instrument approaches and their psychometric properties, themes, types of respondents [29,35,36], and current regulations applicable in Mexico [37–40]. Additionally, the barriers and facilitators involved in promoting healthy eating within schools were explored quantitatively and qualitatively [8]. In the second step, the dimensions of the questionnaire were defined, considering the results of a previous study conducted by our research group, in which parents, teachers and experts responded to a survey and identified seven areas and actions that promote healthy eating as priorities [41]. These components were subsequently organized into the levels of the ecosocial model [42,43], and the definition and importance of the mediators of a SFE for children were considered [28]; teachers were identified as the most relevant respondents for this questionnaire because of the fundamental role they play as mediators of the SFE and as models for their students [44].

In the third step, the characteristics of the target population were considered. For the response scale, a four-level scale was selected to minimize the use of a midpoint [45,46]. As a reference framework, the Centers for Disease Control and Prevention (CDC) School Health Index scale was used, which asks about “actions not implemented”, “actions in the process of being implemented”, “actions partially implemented”, and “actions fully implemented” [36]. At workgroup sessions, the items were selected, considering the themes of the seven dimensions defined, the implementation actions subject to the regulations in force, and the teachers as the respondents. The final wording of the items was to capture the level of implementation of actions, categorized as “none”, “low”, “medium”, and “high”. The name that was chosen for the PWA was NUTRENTO, which and was derived from the prefixes *“nut”, “trend”, and “agent”* to refer to nutrition, trend, and the teacher as an agent of change.

### Content validity

Expert judgment was performed with quantitative assessments using an agreement scale and also with qualitative assessments in the form of suggestions and comments on the items [47]. The items written were sorted by dimension in an Excel sheet and identified with a key according to the abbreviated name of its dimension, the item number within the dimension, and the item number within the questionnaire (S1 Fig).

The number of expert judges was determined based on a minimum sample size of 9 [47,48]. The list of those selected for the JE included professionals with training and postgraduate degrees in nutrition, public policy, health education, physical activity, and psychology; with a line of research and/or field of action in school health; with publications and experience of at least 10 years; with time available to evaluate the questionnaire; and free of conflicts of interest [48]. The invitation to participate was sent by e-mail, and subsequently, the judges who agreed to participate were sent another e-mail with the questionnaire. The judges rated each item concerning its relevance, coherence and clarity, as well as the sufficiency of each dimension. The rating scale was as follows: “Not at all in agreement”, “Slightly in agreement”, “Moderately in agreement”, and “In agreement”. The judges evaluated the items on an Excel sheet that included a space for comments. The evaluation lasted five weeks, and only questionnaires with complete ratings were considered. The responses were integrated into Excel and then analyzed in STATA 18.0. The Aiken V statistic was used to measure the coefficient of agreement between judges. A critical value <0.7 and a merit criterion >0.8 were established [49]. In addition, comments on typographical errors, wording and items suggested for addition by the judges were collected on a similar basis and considered for the corrected version of the questionnaire.

### Progressive Web App development

A framework was followed to develop a PWA [30] according to m-Health principles, which considers self-reports on social interactions with environmental elements, relevance to generate real-time data, adaptation to operating systems to improve accessibility, generation of automatic databases, and massiveness of the scope for a larger target population [26,50,51].

For technological development, React with Typescripty was used as the framework and programming language. As a management tool, JIRA^TM^was used to manage Sprints and development activities, and Bitbucket© was used for code storage and versioning. For access to the web questionnaire, registration was requested by e-mail, with a validation process. In addition, the PWA was required to present a privacy notice and obtain informed consent to collect sociodemographic data (age, sex, rural or urban population, marital status) and teacher profiles (graduate degree, type of contract and working hours) [52]. It also presented the option of keeping the respondent anonymous, voluntarily omitting the sociodemographic and teacher profile data. For the administration of the information generated by the PWA, a content management system (CMS) platform was generated to manage users and download the database, to which only researchers had access.

The perceived functionality of the PWA was tested by applying an instrument that evaluates the usability of Health-ITUES through Google Forms. Responses were obtained from a sample of 16 public elementary school teachers who were recruited through open invitations [53]. The results were satisfactory, with an average of more than 4.5 out of a maximum of 5 points in the dimensions of impact, perceived usefulness, perceived ease of use, and user control [53,54].

### Construct validation

To validate the data obtained from the PWA, exploratory factor analysis (EFA) and confirmatory factor analysis (CFA) were performed on a sample of 970 Mexican teachers. A sample parameter was considered to perform a CFA [55].

### Population and recruitment

A collaboration agreement was reached with the regional education authority to gain access to the schools and carry out the PWA validation study. The study included 244 public elementary schools in 6 municipalities of Hidalgo, Mexico, to which 2644 teachers were assigned. From this population, 123 schools with 1462 teachers were randomly selected. An invitation to participate was addressed to the school principals, who, once informed about the objectives and procedures of the study, determined the feasibility of access to their respective schools. Seventy-six percent of the schools agreed to participate and were visited by a team of professionals trained in the use of the PWA to recruit the relevant users: classroom teachers, physical education teachers, principals and teaching staff with activities associated with the SFE. Our professional staff presented the objectives of the SFE assessment with the PWA to the teachers, requested their consent electronically, and provided technical support and guidance to the participants during the assessment process with the PWA NUTRENTO. The time required for teachers to complete the questionnaire ranged from 2060 minutes, depending on their technological proficiency and the speed of their internet connection. Teachers were not given any incentive and could withdraw from the study at any time. It was verified that teachers could access the domain https://app.nutrento.org/auth/login; additionally, they viewed the privacy notice and registered an email address. Teachers received the validation key in their registered email address to access the PWA. The PWA presented the informed consent form to teachers before they answered the questionnaire. Teachers could answer the questionnaire with the options to provide their identifying sociodemographic data or answer the questionnaire directly, maintaining their anonymity. For the CMS interface, the researchers used the data obtained in the perceived functionality test to check its ability to administer the PWA and verified the automatic generation and downloadability of the database in .cvs format. The evaluations were conducted from 15^th^ February to 29^th^ May 2024 (Fig. 2).

**Fig. 2.**
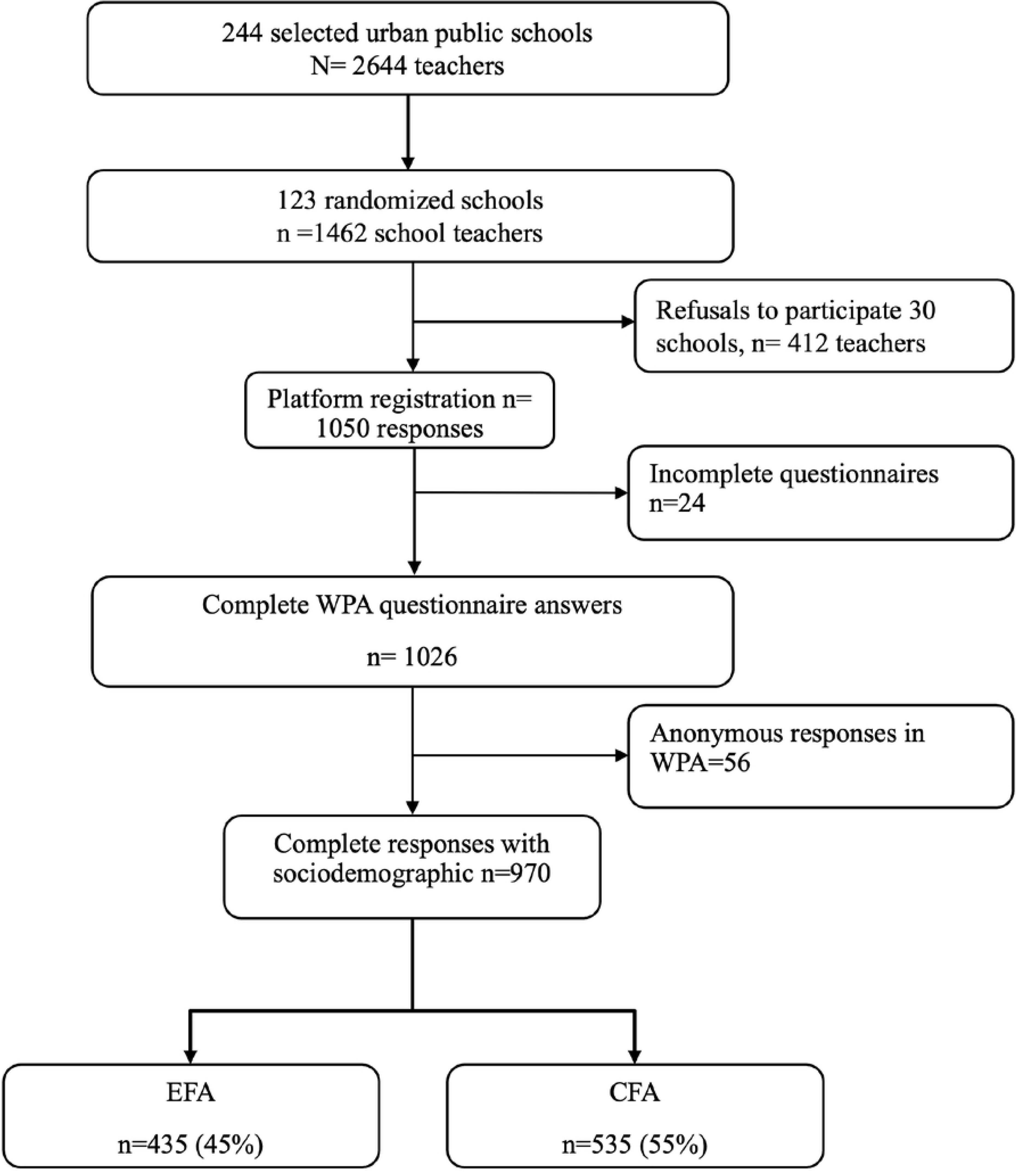
STROBE Flow diagram for the participant population.

After the eligible data were obtained with the PWA, a ratio of 10 was used for the sampling parameter [55], 45% of the sample was randomly selected to perform the EFA, and the rest of the sample (55%) was reserved for the CFA, meeting the criterion for independent sample analysis [56,57].

### Exploratory Factor Analysis

EFA was used to identify the factor structures of the questionnaire [57]. Descriptive tests were performed to determine whether any item should be discarded due to low variability or high skewness. Factor extraction was performed by the common variance method, with varimax rotation to evaluate the internal structure of the items [58]. A categorical response scale was used to determine the relationships among the items first through a polychoric correlation. The relevance of the EFA was assessed by the determination of the matrix correlation (DCM <> 0), the Kaiser‒Meyer‒Olkin test (KMO), and Bartlett’s test of sphericity (> 0.8)[59]. Factors were retained based on eigenvalues >1.0; corrected item–total correlations that were >0.5, and items with communality >0.5 were considered for eliminated. Once factors were eliminated, we proceed to run the AFE again [46,60].

### Confirmatory Factor Analysis

On the basis of the EFA results, models for the CFA were constructed through structural equation modeling with 55% of the remaining sample. A CFA model was estimated by maximum likelihood to obtain a more robust estimate and to standardize it. From this first model, the highest covariances within each factor were identified to rerun the standardized model. To consider a good model fit, the Comparative Fit Index (CFI) and the Tucker‒Lewis Index (TLI) were used with values between 0.90 and 0.95; the Root Mean Squared Error of Approximation (RMSEA) was used for those between 0.05 and 0.08 and p<0.05; and the Standardized Root Mean Squared Residual (SRMR) was used for those p<0.05.

The internal consistency of the items was subsequently evaluated with Cronbach’s alpha, considering adequate values to be between 0.70 and 0.90. Ferbuson’s delta value was estimated for discriminant validity (where 0 indicates “no discriminative ability”, and 1 indicates “completely discriminative”), and Loevinger’s H coefficient (H) was used to evaluate the scalability of the set of items, considering good scalability to be starting from H >0.50, to and poor scalability to be <0.3. Finally, Raykov’s factor reliability coefficient was also determined to determine the composite reliability, with >0.7 as the cutoff point for good reliability [61]. Statistical analyses were performed with Stata 18 for Mac, and where applicable, p<0.05 was considered significant.

### Ethical aspects

The project was approved by the Ethics and Research Committee of the Institute of Health Sciences of the Autonomous University of the State of Hidalgo (folio 200/2023), which is governed by the ethical principles for medical research on human subjects of the Declaration of Helsinki and the Mexican General Health Law [38]. The provisions of the General Law for the Protection of Personal Data in the possession of Obliged Subjects [62] were followed.

The privacy notice on the use and safeguarding of data for research purposes, the guarantee of anonymity and confidentiality, and the informed consent statement were presented to users electronically before they registered for access to the PWA and they were answering the questionnaire at https://app.nutrento.org/auth/register. Respondent acceptance was provided electronically, and in all cases, teacher participation was voluntary. Informed consent was obtained from all participants involved in the study.

## Results

In the conceptualization and literature review stage, the available instruments were identified to assess the SFE properties, which were based on checklists, norms on adherence to the recommendations of food handling and hygiene guidelines, and food guides, as well as the quality of school food services. Twenty-three instruments were identified: 3 (13%) focused on the Mexican context, and only 2 reported face validity and could be answered by personnel outside the school context. We also identified 14 instruments (61%) that had some type of validity; in 13 of them (53%), the respondent was the teacher, and 6 (26%) could be answered online (Table S2). Psychometric limitations and instruments without validation were detected. Similarly, the checklist formats were based on regulations or frameworks with limited relevance or applicability in the Mexican context. Therefore, we proceeded to design a new questionnaire to evaluate the SFE in Mexico, considering the dimensions identified in a previous study conducted by our research group [41], as well as the organizational and educational framework established in 3 general laws [38–40]and 2 regulatory agreements for Mexico [16,63]. The construct “Actions and Elements that promote HSFE” was defined; the instrument considers primary school teachers as respondents so that they can be evaluators and mediators of their own food environment and report information according to the actual school context.

Seven dimensions were defined to evaluate the SFE on the basis of the higher levels of the ecosocial model (community, institutional and organizational), actions and elements associated with financing, school infrastructure, coordination with public institutions, regulation and supervision of standards, and the presence or absence of a school feeding program, and curricular issues associated with nutritional food education (NFE) were identified. At the interpersonal level, implementation items related to the regulation and supervision of normative actions, as well as the perspective and approach to the curriculum related to NFE, were identified (S2 Fig).

In drafting the items, the scope of what schools and school-related processes could implement through teachers was considered. In the first version of the instrument, 47 items were obtained: 7 in the dimension of financial resource management (FN), 8 in the dimension of coordination with institutions (CI), 5 in the dimension of physical infrastructure conditions (IN), 10 in the dimension of the implementation of nutritional food education (NFE), 5 in the dimension of the application of regulations associated with children’s nutritional health (RCC), 4 items in monitoring access to foods high in saturated fat, sugar and salt (MO), and 8 in the implementation of school feeding programs (SFP).

During the content validation stage, 39 judges were identified, 31 of whom agreed to participate, and the questionnaire was sent to them electronically. 22 evaluations were received, of whom 20 were women and 11 were of Mexican nationality. In addition, two judges per country participated from Argentina, Chile, Colombia and Guatemala, and one judge per country participated from Panama, Brazil and Costa Rica. 45% of the judges had doctoral degrees; 86% had backgrounds related to school food and nutrition, 9% in the education sector and 4% in public policy. All the items and dimensions received a favorable evaluation with values above the critical value (V> 0.70), so no item was eliminated. The criterion of clarity was the one that obtained the lowest mean, but not less than 0.7, with the exception of item FN6, which obtained a V of 0.76, which was adjusted to its wording, according to the judges’ comments. The overall V was 0.94, indicating a high agreement coefficient with respect to its proximity to the maximum attainable value of 1 (Table S2). Eleven items that received comments on grammatical connectors were corrected. A corrected version of 52 items of the questionnaire was obtained. As a result of the judges’ suggestions, 5 more items were integrated: 1 item in IN(IN5), 2 in FNE (FNE9, FNE12), 1 in RCC (RCC1) and 1 in SFP (SFP9).

The corrected version of the questionnaire was the basis for the technological development of the Progressive Web App (PWA), which is hosted at the domain https://app.nutrento.org/auth/login. In accordance with the framework proposed for the app, the interface and access were tested so that it could be viewed and accessed from a computer or a cell phone by the user. Fig. 3 illustrates the steps and interface for a mobile device to use the PWA.

**Fig. 3.**
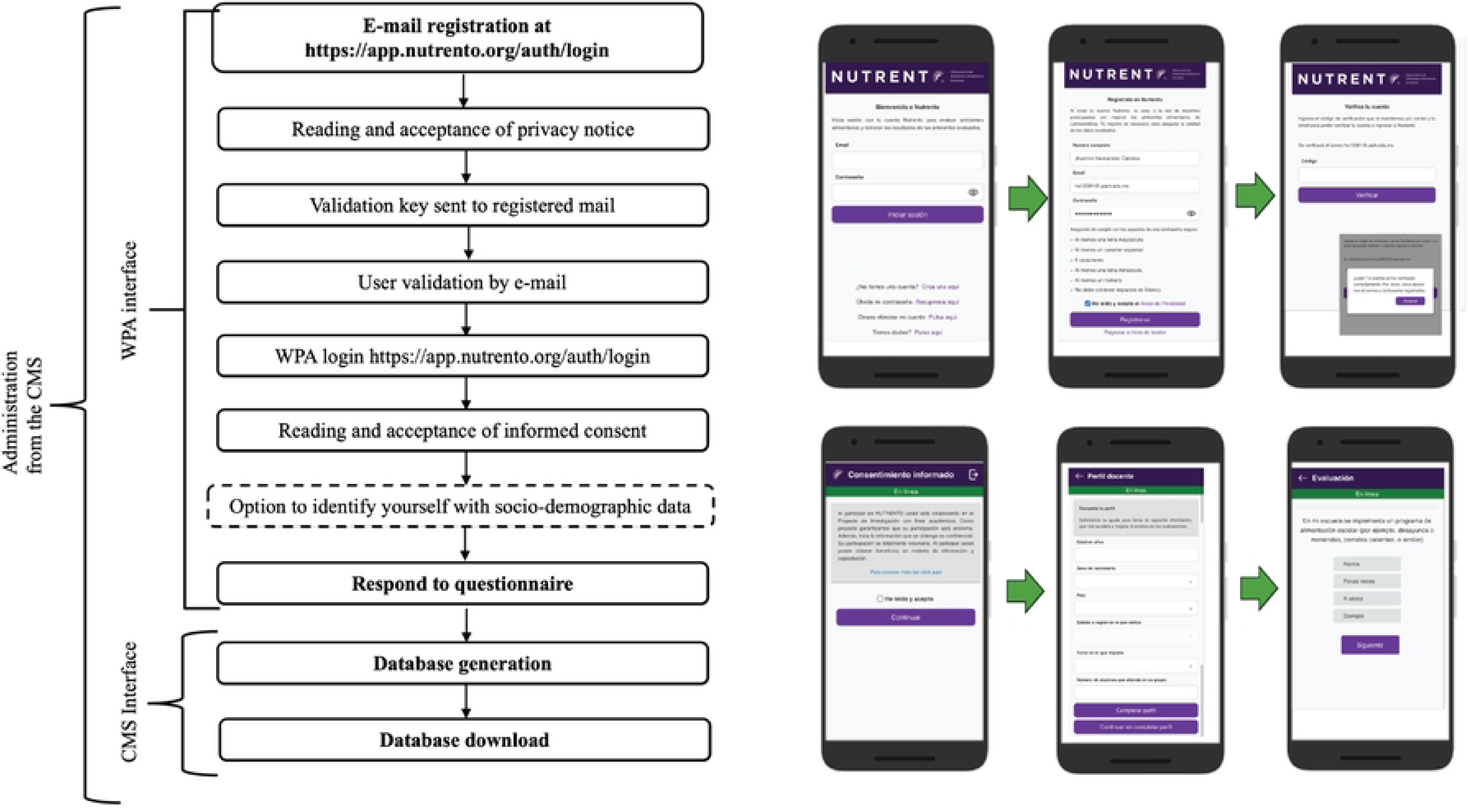
Flowchart with the steps for using the NUTRENTO Progressive Web Application and an example of an interface for a mobile device.

Among the 123 selected schools, 1462 potential respondents to the NUTRENTO PWA questionnaire were identified, of which 30 schools with 412 teachers declined the invitation. A total of 1050 respondents were obtained with the PWA, 24 of whom were eliminated because their responses were not complete. Among the 1026 responses to the PWA questionnaire, 56 (5.5%) respondents decided to keep their responses anonymous (Fig. 2). A final sample of 970 respondents, along with their sociodemographic characteristics and type of employment contract, was obtained and randomized according to the analysis group (exploratory or confirmatory).

The samples used for the EFA and the CFA did not show statistically significant differences in any of the variables, indicating that the randomization process was effective. In both groups, the majority of respondents were women and were between 41 and 50 years of age. Most of them had a basic contract and worked the morning shift (Table 1).

**Table 1.**
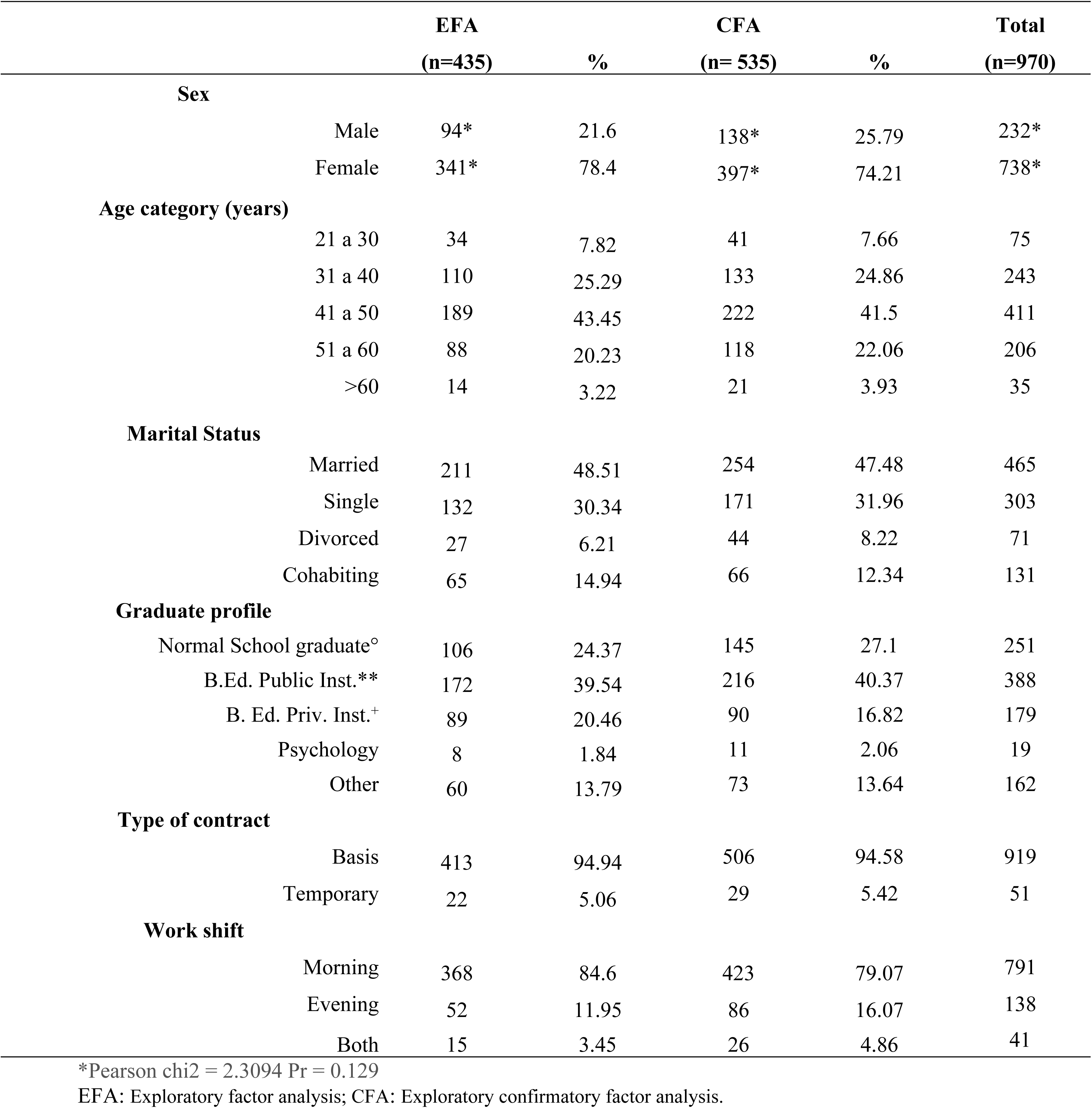
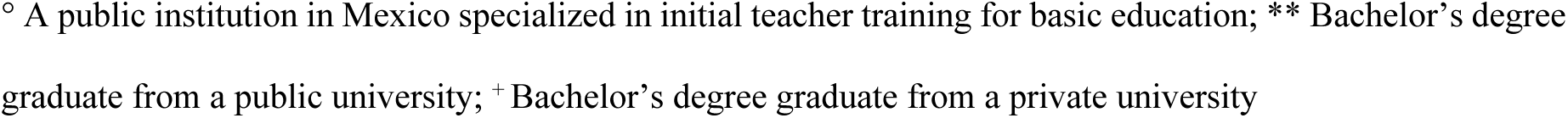
Demographic characteristics of participating teachers from general public elementary schools in 6 municipalities of Hidalgo, Mexico.

Of the sample considered for the EFA of 435 teachers (44.8%), 78.4% were female. Among the total number of respondents included in this analysis, the 41 to 50 year age group had the highest proportion of participants, accounting for 43.4% of the total of this sample. Participants over 60 years of age were the least represented, accounting for only 3.2% of the sample. Among the respondents in this process, 63.9% had teacher training in public institutions (teacher training and a bachelor’s degree in education from a public institution). Nearly 95% of the respondents had a basic contract, 84.6% worked the morning shift, and 3.4% worked double shifts.

For the EFA, 52 items were considered, organized into 7 dimensions. For this first analysis, the DCM was <0.000, and the KMO was 0.94. Bartlett’s test of sphericity yielded a value of Chi-square=14228.119 (p < 0.001). Six factors with eigenvalues greater than 1 were identified, which explained 66.9% of the total variance. Loadings >0.5 were considered, and the subscales showed a factor structure very similar to that proposed in the theoretical model. Factor I grouped the SFP (FI_SFP), factor II the FNE (FII_FNE), and factor III the monitoring (FIII_MO). Factor IV grouped the CI items (FIV_CI), V the FN (FV_FN) and MO the RCC (FVI_RCC). Items with uniqueness >0.70 were eliminated. SFP and MO did not affect the internal structure. For FNE, items FNE1, FNE2, FNE3, FNE4 and FNE9 were removed. For CI, items CI5 and CI6 were removed, and for FN, items FN2 and FN4 were removed. For RCC, RCC4, RCC5 and RCC6 were removed. Only item IN3 of the IN subscale was grouped into factor VI (Table 2).

**Table 2.**
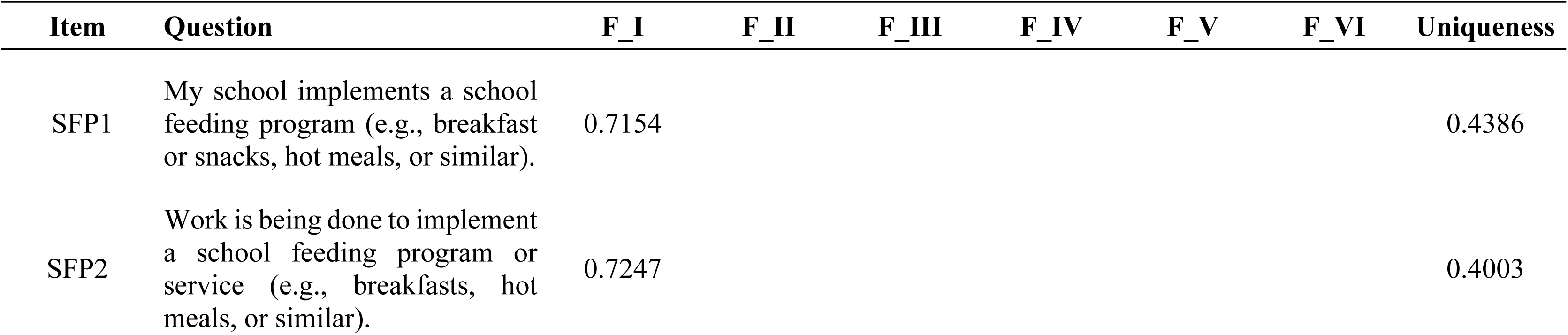
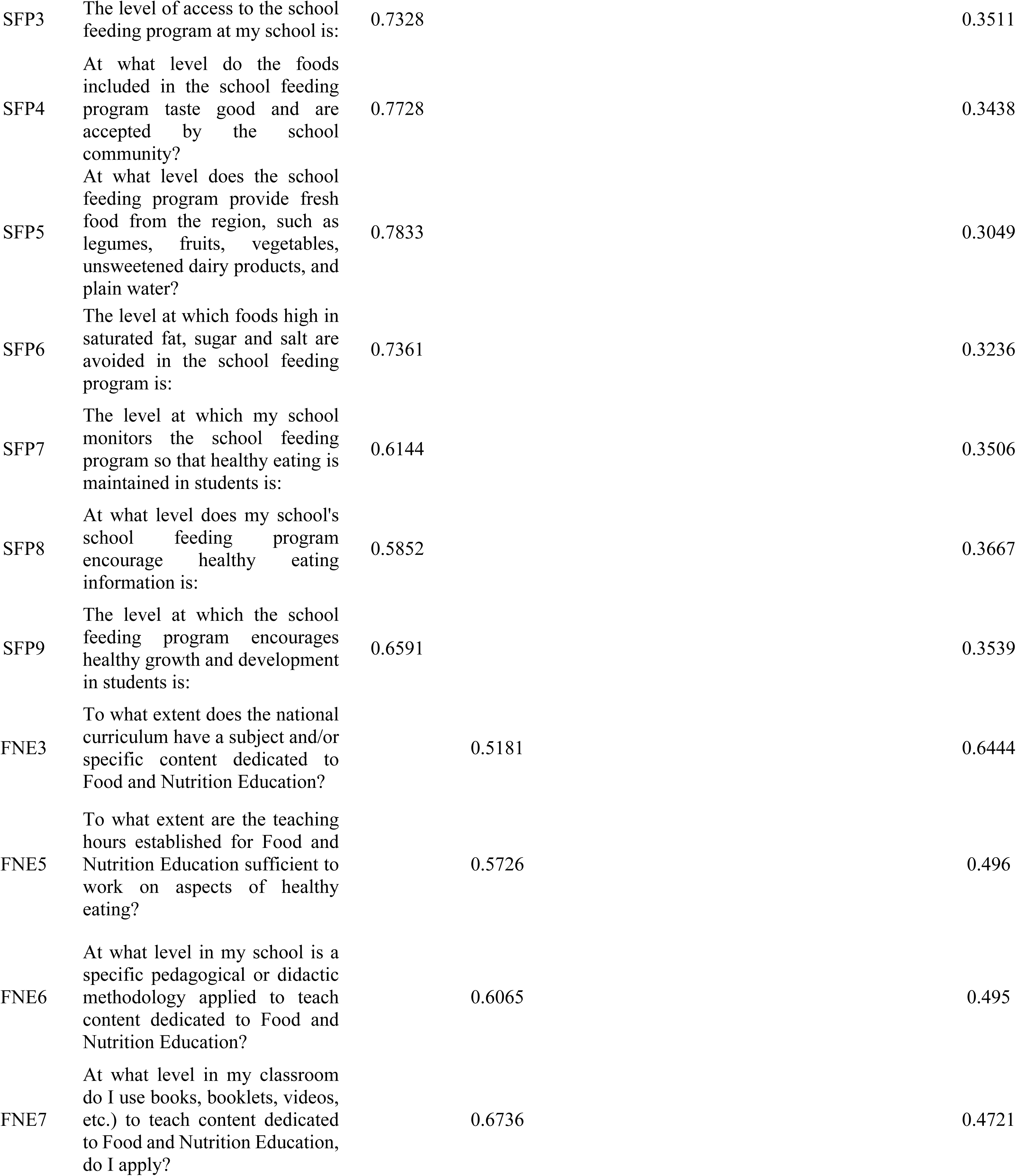
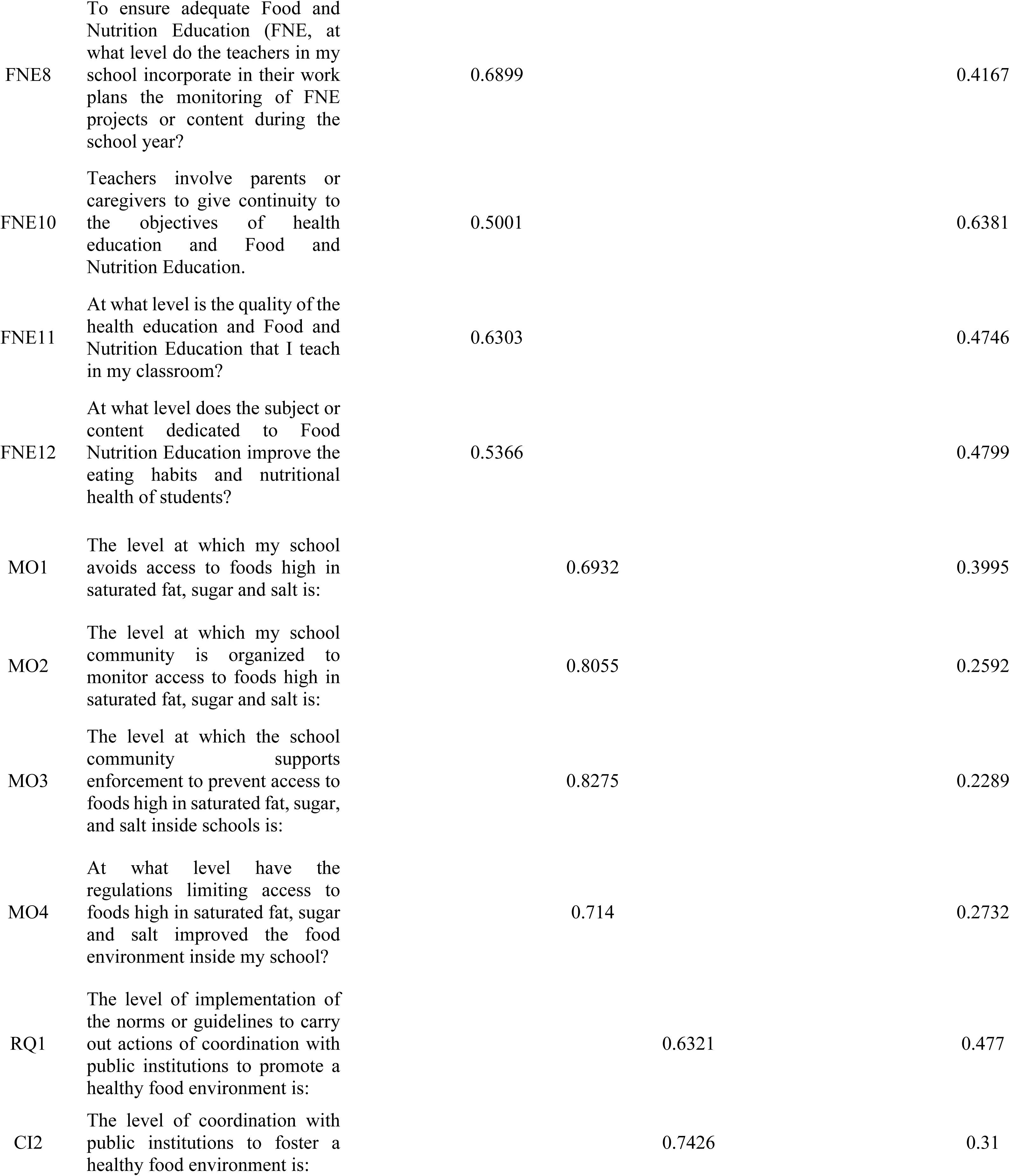
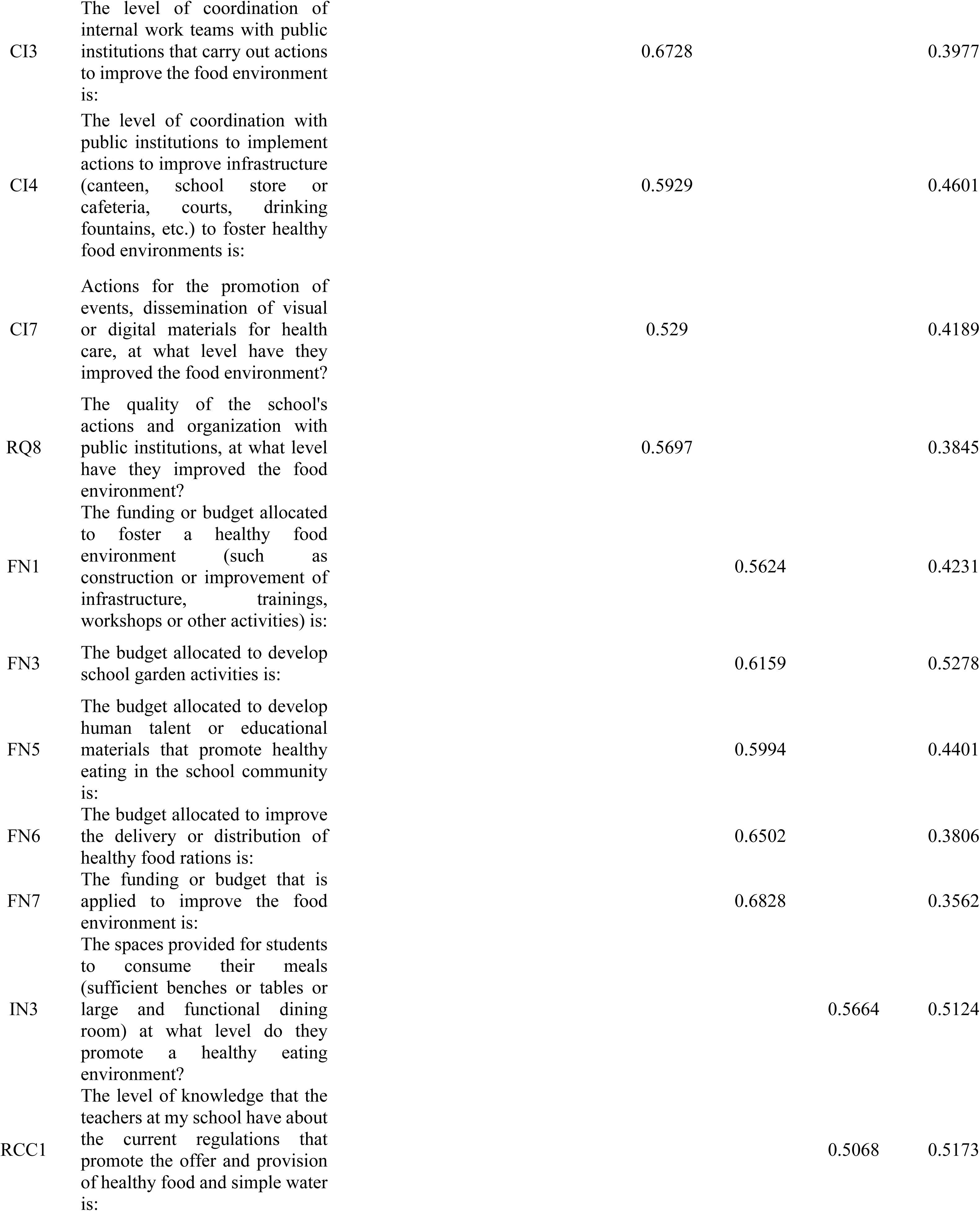
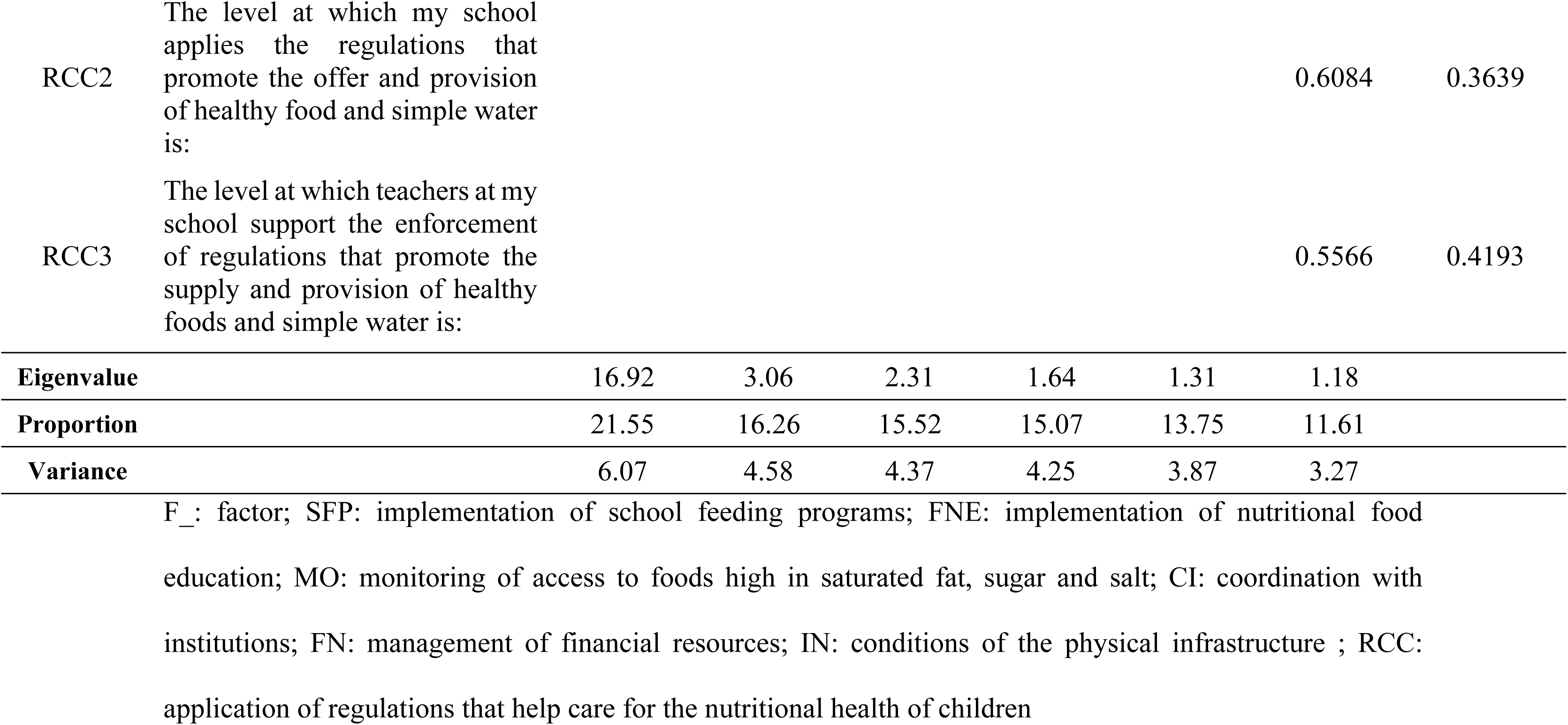
Final structure of factors identified for the Progressive Web Application NUTRENTO questionnaire.

Only 36 items were considered for the AFC. In the confirmatory model for six factors, when the fit of the model was evaluated compared with that of the null model, the CFI was excellent; similarly, the TLI value was at the upper limit of good fit, which was considered adequate, at 0.945. The RMSEA was an excellent fit at 0.045, and for the standardized difference between the observed and predicted correlations, the SRMR was excellent, at 0.059 (Table 3). Fig. 4 shows the conformed factors with their respective items and their standardized coefficients. The components analyzed showed consistency in their factorial weights for each of the 36 items grouped into six factors. The relationships between the factors were significant, and their standardized regression coefficients were greater than 0.30, in a range of 0.50 to 0.91. The FI_SFP was formed with the original theoretical scale, with factor weights ranging from 0.67 (for SFP1) to a maximum of 0.83 (for SFP3). The FII_FNE was also formed congruently, with factor weights between 0.51 (for FNE3) and 0.78 (for FNE8).

**Figure 4.**
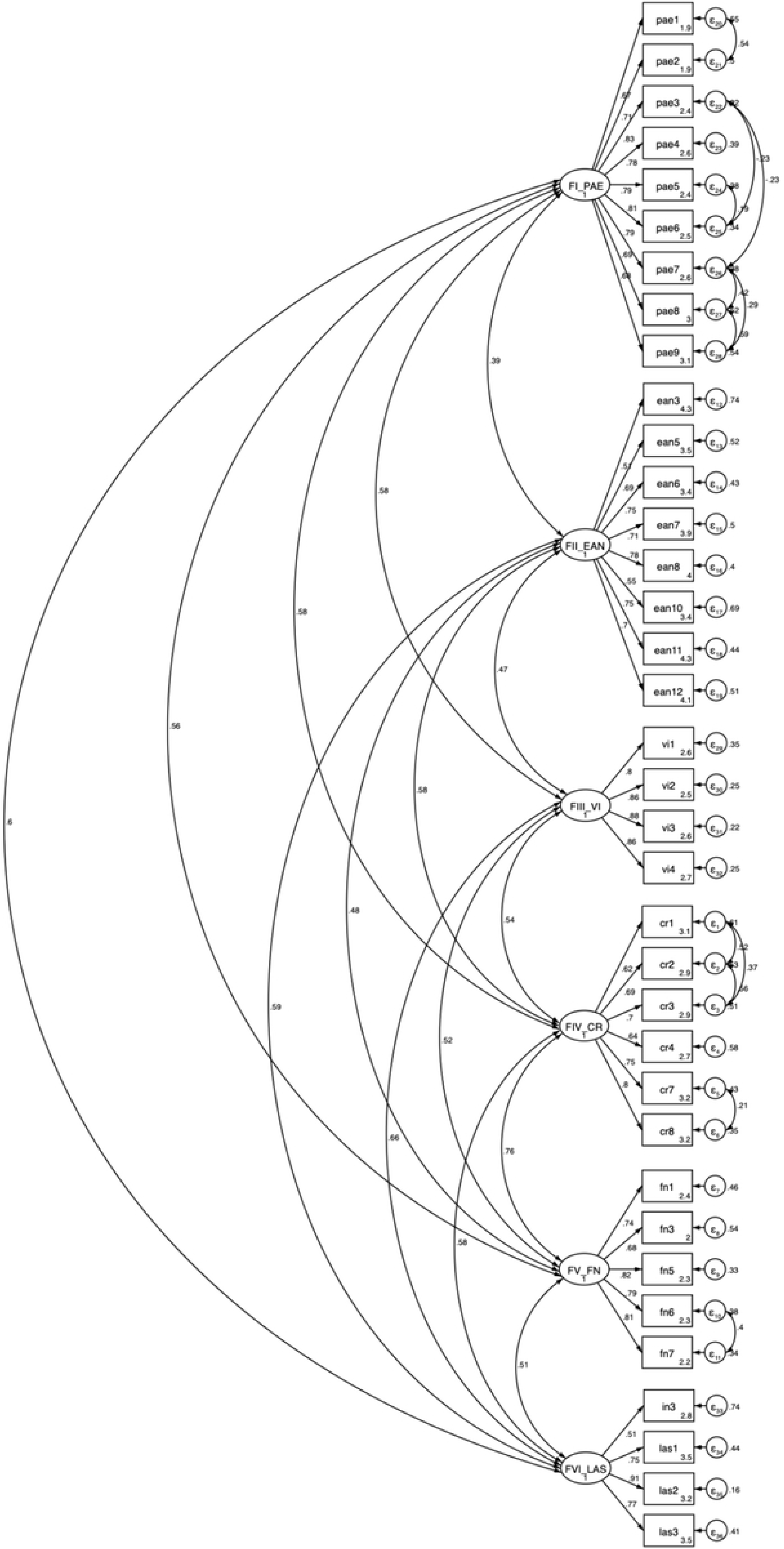
Six-factor model of the questionnaire to identify the levels of actions and elements that foster a healthy school food environment in the NUTRENTO Progressive Web Application.

**Table 3.** Statistics of the model with CFA fit indices to the questionnaire for the Progressive Web App NUTRENTO.

|  | Acceptable<br>fit index<br>criterion | Base Model | Adjusted<br>model |
| --- | --- | --- | --- |
| **CFI** | > 0.90 | 0.887 | 0.95 |
| **TLI** | > 0.90 | 0.877 | 0.945 |
| **RMSEA** | <0.80 | 0.067 | 0.045 |
| **SRMR** | <0.80 | 0.066 | 0.059 |

CFI: comparative fit index; TLI: Tucker-Lewis index; RMSEA: root mean squared error of approximation
SRMR: standardized root mean squared residual.

For FIII_VI, the factor loadings of the model were greater than 0.8 for all the items. For FIV_CI, the highest factor loading was 0.8 (for CI8), and the lowest was 0.62 (item CI1). For FV_FN, the lowest loading was 0.68 (for FN3), and the highest loading was 0.82 (for FN5). Finally, FVI_RCC had the lowest loading, 0.51 (for IN3), and the highest, 0.91 (for RCC2).

The internal structure of the confirmed model with six factors in this research is supported by the significant factorial saturations (p < 0.001) between the factors and between the factors and the items.

After the CFA, the internal consistency of the factors obtained was evaluated by Cronbach’s alpha. The alpha was adequate for all the factors (greater than 0.80 and less than 0.95); FI_SFP was the factor that showed the highest consistency (alpha of 0.92), and F_VI_RCC was the one with the lowest but acceptable consistency (0.82). The individual consistency of the items was also tested. Loevinger’s H coefficient was greater than 0.5 for all factors in the range of 0.55 and 0.78 for FIII_FNE and F_VIRCC, respectively (Table 4). The composite reliability according to Raykov’s factor reliability coefficient of all factors was greater than 0.7. When determining the alpha of the items, for factor six, RCC1, RCC2 and RCC3 had alphas of 0.76, 0.7 and 0.75, respectively. These items were retained since the overall alpha of the subscale was 0.82. Additionally, the H of the items was >0.5, and the overall alpha was 0.6.

**Table 4.** Item and factor reliability were determined by Cronbach’s alpha and Loevinge’s H coefficient, and composite reliability was determined by Raykov’s factor reliability coefficient of the questionnaire for the Progressive Web App NUTRENTO.

| FI_SFP | $\alpha$ of item | $\alpha$ of the factors | Item H | H factor | Raykov's FC |
| --- | --- | --- | --- | --- | --- |
| SFP1 | 0.92 | 0.92 | 0.61 | 0.64 | 0.889 |
| SFP4 | 0.92 |  | 0.65 |  |  |
| SFP5 | 0.91 |  | 0.66 |  |  |
| SFP3 | 0.92 |  | 0.64 |  |  |
| SFP6 | 0.91 |  | 0.65 |  |  |
| SFP7 | 0.91 |  | 0.65 |  |  |
| SFP9 | 0.91 |  | 0.64 |  |  |
| SFP8 | 0.92 |  | 0.64 |  |  |
| FII_FNE |  |  |  |  |  |
| FNE3 | 0.87 | 0.87 | 0.48 | 0.55 | 0.872 |
| FNE5 | 0.86 |  | 0.59 |  |  |
| FNE6 | 0.85 |  | 0.59 |  |  |
| FNE7 | 0.85 |  | 0.56 |  |  |
| FNE8 | 0.85 |  | 0.59 |  |  |
| FNE10 | 0.87 |  | 0.45 |  |  |
| FNE11 | 0.85 |  | 0.59 |  |  |
| FNE12 | 0.86 |  | 0.54 |  |  |
| FIII_VI |  |  |  |  |  |
| VI1 | 0.9 | 0.91 | 0.77 | 0.78 | 0.914 |
| VI2 | 0.88 |  | 0.79 |  |  |
| VI3 | 0.88 |  | 0.79 |  |  |
| VI4 | 0.89 |  | 0.77 |  |  |
| FIV_CI |  |  |  |  |  |
| CI1 | 0.86 | 0.88 | 0.61 | 0.6 | 0.783 |
| CI2 | 0.85 |  | 0.65 |  |  |
| CI3 | 0.85 |  | 0.64 |  |  |
| CI4 | 0.88 |  | 0.54 |  |  |
| CI7 | 0.87 |  | 0.56 |  |  |
| CI8 | 0.86 |  | 0.59 |  |  |
| FV_FN |  |  |  |  |  |
| FN1 | 0.88 | 0.88 | 0.61 | 0.68 | 0.864 |
| FN3 | 0.88 |  | 0.68 |  |  |
| FN5 | 0.85 |  | 0.68 |  |  |
| FN6 | 0.85 |  | 0.69 |  |  |
| FN7 | 0.84 |  | 0.72 |  |  |
| **FVI_RCCIN** |  |  |  |  |  |
| IN3 | 0.85 | 0.82 | 0.49 | 0.6 | 0.829 |
| RCC1 | 0.76 |  | 0.59 |  |  |
| RCC2 | 0.7 |  | 0.68 |  |  |
| RCC3 | 0.75 |  | 0.62 |  |  |

$\alpha$ Chronbach's Alpha; H: Loevinger coefficient.
SFP: implementation of school feeding programs; FNE: implementation of nutritional food education; VI: monitoring of access to foods high in saturated fat, sugar and salt; CI: coordination with institutions; FN: management of financial resources; IN: physical infrastructure conditions; RCC: application of regulations that help care for the nutritional health of children.

## Discussion

The objective of this work was to develop and validate a progressive web application aimed at primary school teachers to identify the levels of actions and elements that promote healthy school food environments in Mexico.

For the evaluation of the school food environment, there is no single unit of measurement that is specifically for this purpose [25]; however, the proposal of this work from the conceptualization phase focused on the approach of the construct on the level of actions or elements that promote an HSFE. Schools, owing to their normative frameworks, are already carrying out some types of actions [16,40,63]. These actions could be implemented by teachers as mediators of HSFEs, so the dimensions addressed aspects of the implementation of the school feeding program, surveillance, nutritional food education, coordination with institutions, and the search for funding and implementation of regulations for the nutritional health care of schoolchildren.

In a review of the literature, no tool adapted to the context of elementary schools in Mexico was found. The instruments that were identified focus on physical attributes, and although they touch on regulatory or public policy aspects, very few instruments include procedural or implementation aspects that have social implications, such as the perspective of those who implement the regulations, prioritization issues of schools, and the presence or absence of teaching methodologies or environmental support for food and nutrition education carried out in schools [25]. Therefore, a proposal focused on the topics prioritized by the school community involving the promotion of an HSFE was developed [41]. In addition, the development of the construct “Actions and Elements that promote HSFE” implied a more holistic approach, as seen in designs such as the one made by Moore and collaborators, which also recognizes the value of the involvement of SFE mediators in the different processes to implement actions that are expected to result in an HSFE [64]. In this research, the construct considered issues of more distal dimensions (such as coordination with institutions and financing) that support the more proximal dimensions (such as the implementation of the school feeding program, nutritional food education, monitoring, and the implementation of regulations). The above is relevant to the Mexican context, since the models for implementing policies and operational regulations associated with food environments fall under the autonomy of schools [39,63]. Although there are proposed instruments to measure SFE, such as the SEAT, which focuses on availability [65], or others reported by Saluja [22], which complement elements of the school canteen, diet, and physical activity of students, they do not correspond to the context for which this instrument was proposed. Therefore, an instrument that explores the underlying factors and that measures availability could be more relevant for the Mexican and Latin American context. Other instruments include items available for the promotion of healthy habits; however, including only these items could lead schools to prioritize other activities or to perceive that the SFE is configured with a short promotional action, with only providing information, or addressing curricular content in isolation [66].

The PWA approach is relevant to current needs, as m-Health has gained relevance because it is increasingly present in different areas of public health [67]. The use of these digital tools for the SFE could be considered a new strategy and tool to monitor and prevent childhood obesity [26]. In addition, few published electronic instruments exist, and those that exist were developed in the context of high-income countries [68] or were limited to aspects such as school lunch [69] or evaluations of the quality of food service [44,70–72].

In other studies where digital tools have been developed, such as those conducted by Morin in 2021 and by Patterson [70] and Marty in 2024 [73], the validation of content and psychometric properties has been based on interrater reliability. However, our study focused on the confirmation of the conceptual model, and all the steps for the validation of an instrument were carried out [61,74], starting from an EFA to visualize the internal theoretical structure proposed from the theoretical model, through which the number of items could be reduced.

The EFA showed very meritorious indicators, both for the DCM (p<0.001), a KMO of 0.95, and a Bartlett test of sphericity (p<0.001). This was the case for the primary EFA with 52 items (Table S3), as well as for the final EFA, which resulted in a shorter questionnaire of 36 items. By reducing the number of items by 30% with respect to the original questionnaire, the time investment for using this application could be minimized in the future, as the instrument is more parsimonious. [74]. For the primary EFA, items with uniqueness greater than 0.7 were eliminated (FNE1, FNE2, FNE3, FNE4, and FNE9; CI5 and CI6; FN2, FN4; RCC4, RCC5, and RCC6; IN1, IN2, IN4, and IN5). These eliminations were discussed with the research team and was owing to the possible redundancy of the questions [46,60,74]. This also implied the elimination of 4 out of 5 items of the subscale, which was theoretically proposed for school infrastructure (IN). The plausibility of the items within the theoretical structure and their operation within the schools could have been affected by external elements of the schools beyond the scope of the mediated action of the teachers; alternatively, as in the case of the items in FNE1, (on the responsibility that the teacher perceives to have for the nutritional food education of their students), there are other intrinsic elements of personality or teacher training, which could be influencing factors. Item IN3 is associated with a question about the infrastructure for food consumption: “The spaces provided for students to consume their food (sufficient benches or tables or large and functional dining room) at what level promote a healthy eating environment?”); was grouped with a loading of 0.56 in the factor VI_RCC, and since it was the only item associated with infrastructure that was considered plausible and for which teachers could report their assessments, it was integrated into the factor FVI. The EFA with 36 items resulted in more acceptable factor loadings greater than 0.5. On the other hand, the elimination of the items improved the uniqueness with respect to the primary EFA (Table S3).

The adjusted and standardized model for the CFA showed acceptable higher correlations at > 0.3 for all factors. Similarly, the goodness-of-fit indices CFI, TLI, RMSEA, and SRMR were excellent and adequate [46,60,75], with values of 0.95, 0.45, 0.045, and 0.059, respectively. The reliability of the model was high. FI_SFP had alphas greater than 0.90, whereas FII FNE, FIII VI, F IV CI, and FV FN had reliabilities between 0.85 and 0.90. Although factor six presented alpha values of 0.7 for items RCC1, RCC2 and RCC3, this level is considered acceptable in the literature. Similarly, the FVI factor presented an overall alpha of 0.82. Therefore, both the model and the items showed high reliability [76].

The dimensions that make up the SFP instrument, RCC and VI, are similar to those found in other instruments that consider the evaluation of SFE [29]. These dimensions are directly associated with policies that shed light on food regulations, mainly to promote healthier eating and restrict access to undesirable foods. However, implementation in Mexico and Latin America is a challenge. School policies associated with preventing or reducing the consequences of chronic noncommunicable diseases have shown potential effectiveness. However, these evaluations have focused mainly on high-income countries, where infrastructure, school feeding programs, and technological and administrative elements for surveillance could receive more funding for implementation. In addition, few studies have evaluated the impact of such policies on any indicator of nutritional status [77].

One of the main strengths of this study is the validation of the “Level of actions that promote healthy school environments” construct. This was developed from a solid conceptual design on the basis of a literature review—which, for the most part, did not find evidence for a contextualized approach in Mexico and Latin America—and background information that allowed us to prioritize the key issues and establish the basis for the adequate operationalization of the construct.

Another relevant strength is the relevance of the study for the analysis and strengthening of the SFE in the Mexican context. The construct was carefully designed following methodological guidelines and recommendations for the development of valid and reliable instruments. This methodological rigor supports the reliability of the questionnaire and provides an accurate assessment of the level of implementation of actions aimed at promoting healthy school environments. Unlike other instruments that focus mainly on physical aspects, inventories or food availability, the PWA NUTRENTO emphasizes the role of the teacher as the central mediator of such actions within the school.

The aforementioned tool also made it possible to explore teachers’ responses within the environmental context implied by the questionnaire. The responses of the teachers, with respect to the scope of m-Health, EMA theories, may be promising for the initiation of actions. Also, this work presents a web instrument with free access that may be accessed from any type of device without being limited by specific operating systems, enables the ability to obtain responses in real time, considers a focus beyond the food available, includes the mediating actor of the school food environment, and explores the level of implementation of the actions that may or may not be being carried out in the school. Qualitative studies have reported that teachers assume responsibility and are important agents in food and nutrition education and that their actions can have a positive impact on their students [11,78]. Therefore, self-reporting tools should be explored as a relevant means to evaluate and plan actions in the school food environment.

The results of the construct validation of this research provided an instrument with validity and reliability to enable teachers in Mexican contexts to evaluate the level of actions and elements within almost 100,000 schools in Mexico and promote healthy school environments. In schools in Mexico and Latin American countries, school feeding programs are not universal, and the food supply is often unhealthy, as has been reported in local studies, which have discovered that most schools do not comply with the regulations [79–81]. Consequently, having only one tool that inventories the type and availability of food would be short-sighted.

Among the limitations of this study, it should be noted that interrater reliability was not considered for the instrument. In particular, the teacher’s perception of his or her reality within the classroom and school may be affected because he or she is the main mediator of the school food environment and performs actions commissioned by his school authority, and what he or she has experienced and implemented is different than that of his or her peers, which may influence his or her answers. Future research should investigate the degree of reliability between the responses of teachers from the same school.

Another limitation of this study is that the teachers answered the questionnaire in group meetings in which school authorities participated, so they may have provided favorable answers due to social desirability or positive or negative motivations [82].

Additionally, the instrument does not include specific questions about actions or elements related to physical activity. This dimension is of utmost importance, so it may be necessary to design a toolkit to measure healthy school environments that includes physical activity and other movement behaviors [65].

Owing to the complexity of school food environments, a toolkit could be required that, in addition to addressing food elements, focuses on different respondents, such as parents, school and health authorities, and researchers, with the aim of integrating actors who can perform assessments that allow synergistic actions in different areas involving SFE [65].

## Conclusion

This tool represents a significant advance by offering, for the first time in Mexico, a progressive web application with a validated digital instrument that allows the evaluation of the level of implementation of actions for a healthy school food environment from the teacher’s perspective. Its accessibility from any device with an internet connection makes it a scalable option and easy to adopt in diverse school contexts. However, it is necessary to continue evaluating its applicability in different educational environments and its usefulness to guide the design and implementation of more effective and sustainable interventions.

## Data Availability

The data presented in this study are not publicly available due to privacy restrictions. Data are available on request from the corresponding author.

https://nutrento.org/ligas-de-interes

## Acknowledgments

The authors thank the Instituto Hidalguense de Educación for its collaboration, and the teachers for their participation in the different stages of the project. Special thanks to the researchers and field team of the Nutritional Epidemiology Research Group (CAEPINUT) of the UAEH.

## Supporting Information Citations

**S1 Fig. Excel Matrix for Evaluating the Content of the Questionnaire for the Progressive Web Application NUTRENTO**

**S2 Fig. Organization of the questionnaire dimensions for the NUTRENTO Progressive Web Application, according to the ecosocial model**

**S1 Table. Instruments identified for the assessment of the school food environment**

**S2 Table. Average Aiken V (V) for items in terms of clarity, consistency, and relevance**

**S3 Table. Primary exploratory factor analysis with seven factors corresponding to the NUTRENTO Progressive Web Application questionnaire.**

